# Individual’s daily behaviour and intergenerational mixing in different social contexts of Kenya

**DOI:** 10.1101/2021.03.10.21253281

**Authors:** Emanuele Del Fava, Irene Adema, Moses C. Kiti, Piero Poletti, Stefano Merler, D. James Nokes, Piero Manfredi, Alessia Melegaro

## Abstract

Which are the characteristics of contact patterns in diverse social contexts in sub-Saharan Africa, and which types of individuals and daily behaviours may play a pivotal role in infection transmission to the most vulnerable, such as older adults? We address these questions using novel survey data on social contacts and time use from a sample of 1407 individuals from rural, urban, and slum settings in Kenya. In the rural setting, we observed the highest number of daily social contacts (11.56, SD = 0.23) and the highest share of intergenerational mixing with older adults (7.5% vs. around 4% in the urban settings). Intergenerational mixing with older adults was mainly reported by individuals spending their day mostly in the general community (around 8%) or at home (5.1%), rather than at work (1.5%) or at school (3.6%). These results are essential to define effective interventions to control infection transmission in the African context.

---

Individual social interactions are critical drivers of transmission processes underlying the spread of infectious diseases such as measles, influenza, and the novel coronavirus infection (SARS-CoV-2). The analysis of age-specific mixing patterns is key to identifying the main transmission routes bringing the disease into the most vulnerable population segments. This becomes crucial when considering resource-poor settings, where the quality and availability of health care requires special efforts to define feasible and sustainable strategies to prevent transmission and reduce the disease burden.

Evidence from social contact surveys has shown that, in low- and middle-income countries (LMICs), contact patterns are generally characterized by (i) a marked age assortativeness (i.e., individuals tend to interact more often with peers of similar age) concentrated among school-age children and young adults, and (ii) higher rates of intergenerational mixing compared to high-income settings. Common features from previous studies are that the largest daily number of contacts is experienced by school-aged children between 6 and 18 years old and working-age adults, while intergenerational contacts are mainly reported between household members^1–11^.

The heterogeneity in the observed contact patterns appears to be strongly related to the social context in which these surveys were conducted. These include the geographical area – with studies being carried out in sub-Saharan Africa (Kenya, South Africa, Uganda, Zambia, and Zimbabwe), East Asia and Pacific (China, Fiji, Thailand, and Vietnam), and Latin America (Peru); the urbanization level – with conflicting evidence for the difference in contact numbers between rural and urban settings^5,7,8^; the spatial distribution – with age-assortativeness and distance from home being positively correlated^8^; the activity status of individuals – with the number of contacts reported in a given location being positively correlated to the time spent in that location^7^; the ethnicity – with minimal mixing found between people of different ethnic background ^10^.

These findings remark the importance of gathering further data to shed light on the differences in mixing patterns between different social contexts, especially in countries characterized by large socio-economic differentials at varying levels of urbanization. Indeed, individuals living in diverse settings along the rural-urban gradient are shown to suffer diverging morbidity and mortality paths because of the interplay of environmental and socio-economic factors^12–15^.

The quantification of social contact patterns has led to the development of more refined computational models to evaluate public health policies aimed at preventing, containing and mitigating the disease spread, at defining appropriate vaccination campaigns or at designing and projecting the impact of non-pharmaceutical interventions (NPIs) as case isolation, social distancing, school closures or reduction of economic activities ^16–19^.

The clinical outcomes experienced at different ages of infection and the role of different age groups in shaping the transmission strongly depend on the infection considered. For instance, infections by respiratory syncytial virus (RSV) mainly affect children below five years of age ^20^, with school-age children as the most likely source of infection for infants, therefore both representing the best targets of vaccination campaigns ^21,22^. Conversely, evidence from the current COVID-19 pandemic strongly suggests that older adults, along with people affected by chronic conditions, are at higher risk of experiencing severe disease and death^23–26^, while children are less susceptible to the infection and young adults are considered silent spreaders as they are more likely to be asymptomatic or paucisymptomatic compared to older ages^27,28^. To add to the clinical challenges, also the contextual complexities, especially in LMICs, have to be considered where the huge heterogeneity characterizing people mobility and access to work opportunities, and the daily routine of individuals living in different socio-economic conditions are critical determinants of the epidemiological outcomes ^7,15,19^.

For this purpose, we conducted a population-based survey in three diverse settings on the southern coast of Kenya and gathered data on social contact patterns and individuals’ daily routines from a sample of 1407 individuals. We sought to assess whether differences existed between urban and rural settings and how these differences affected age-specific social mixing patterns. To this aim, study sites were selected to emphasize differences in terms of urbanization and living conditions. In particular, our study population consisted of (i) one rural village within the Kilifi Health and Demographic Surveillance System (KHDSS) in Kilifi county ^29^, and (ii) two urban areas in Mombasa county, the second largest Kenyan city, namely, an informal settlement (“slum”) and an urban mixed area, characterized by a mix of slum and non-slum elements.

As part of the study, participants reported the number of different persons encountered in two randomly assigned consecutive days, the type and the location of such interactions, the age of contacted individuals, and the socio-demographic characteristics of households, schools, and workplaces. From the same individuals, we also collected time-use information on daily routines, quantifying the proportion of time spent during each day at home, school, work, and in the general community.

Our study innovates in two main directions: (i) we performed a comparison of social contact and mixing patterns in three extremely different socio-demographic settings within a LMIC; (ii) we identified which groups of subjects and behaviours can play a pivotal role in infection transmission across different age groups, as a consequence of the interplay of their age mixing and daily patterns. Additionally, since our data were collected before the COVID-19 pandemic in Kenya, we furnished a valuable benchmark to assess the impact of NPIs on the COVID-19 spread and to evaluate possible vaccination strategies in the country. As the three settings considered in our study cover a wide part of the rural-urban gradient, they may be also used to evaluate control policies in those sub-Saharan countries for which contact data are still lacking.

## Results

### Socio-demographic characteristics of the study population

The sample included 1407 individuals, who filled in a contact diary during two different days. As some participants dropped out after the first survey day, we ended up with a total of 2705 person-days (p.d.) observed. The characteristics of survey’s participants and their mean reported contacts are summarized in Table 1.

**Table 1.**
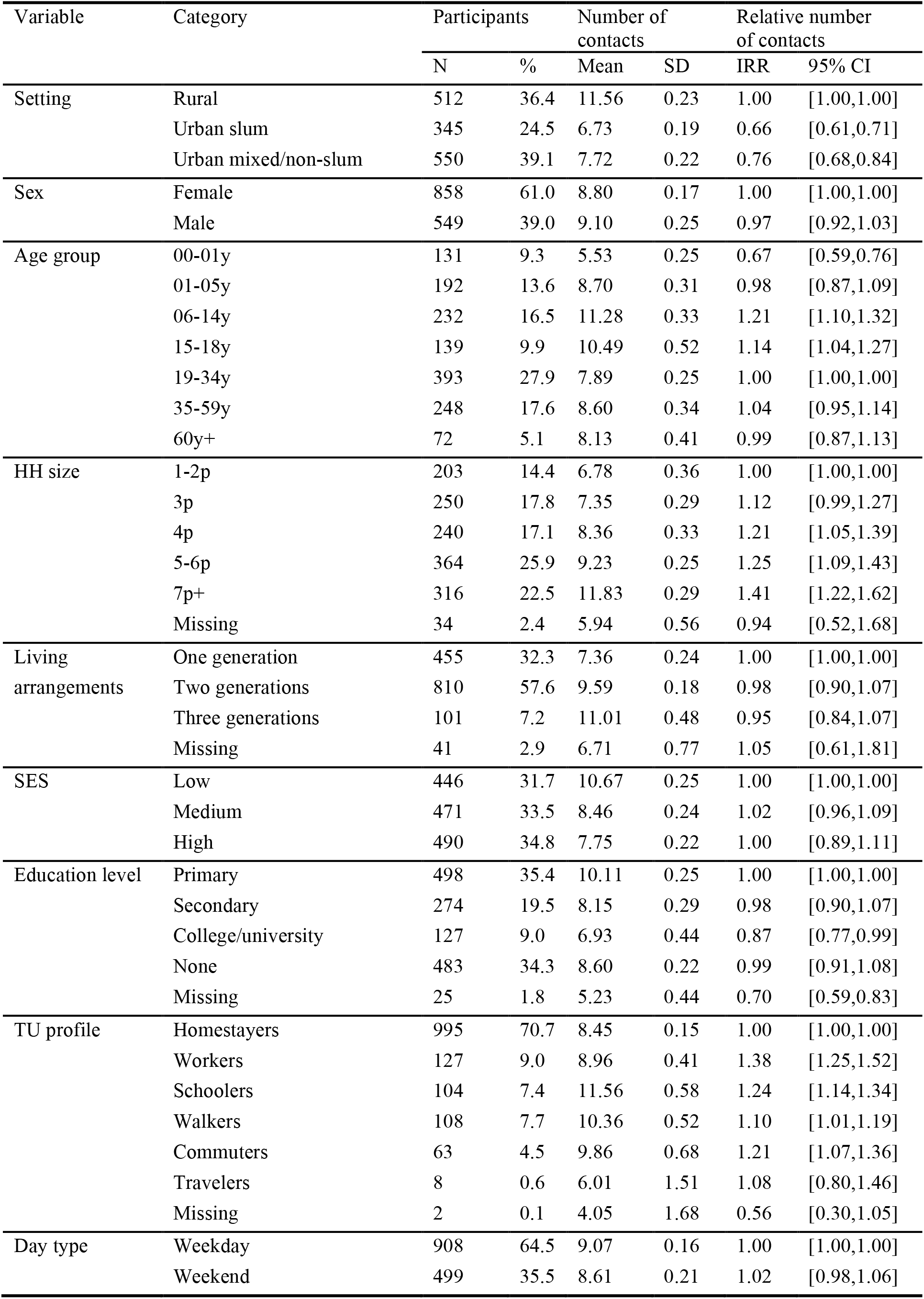
Characteristics of participants and their daily contacts. The relative numbers of daily contacts (incidence rate ratio, IRR) was obtained from a GEE model with negative binomial distribution and overdispersion parameter equal to 0.14, 95% CI [0.13, 0.16].

To assess the characteristics of widely diverse settings in terms of socio-economic status (SES), we compared the rural setting (512 individuals, 1019 p.d.), which presented the typical young age structure of a growing population, with the two urban settings, namely, the slum (345 individuals, 645 p.d.), and the combined mixed and non-slum setting (550 individuals, 1041 p.d.), which were characterized by a lower proportion of young people and a larger proportion of people in the work-age category (Supplementary Fig. S1), as expected for a population experiencing different stages of the demographic transition^30^.

After adjusting for survey design with sampling weights ^7^, relevant differences between settings were found in terms of sex, age, household (HH) size, SES, and type of survey day (Fig. S2 in the supplementary material). Specifically, people living in the rural setting were on average younger, lived in larger HHs, and had a lower socio-economic status, both in terms of SES index and current education level.

### Social contact patterns by setting

A total of 23,532 contacts were reported over the two survey days. The overall mean number of contacts per person per day was 8.92 (median 8, IQR 5-12) and it was higher in the rural setting (mean 11.56) than in the two urban ones. After adjusting for the other participants’ characteristics, we found that the average number of contacts was 34% and 24% lower in the slum and the mixed urban area, respectively (Table 1).

For each of the three settings, we built contact matrices by five-year age groups defining the intensity of interactions between the age groups (Fig. 1). For each matrix, we calculated the assortativity index Q, which quantified the importance of contacts occurring between individuals of the same age^31–33^. The lower the value of *Q*, the lower the assortativity and the higher the mixing between different age groups (Fig. 1). In each setting, we found predominantly age-assortative contact patterns among younger individuals, with a larger number of contacts occurring among school-age children and adults up to 30 years. Overall, the index *Q* was equal to 0.061 for the rural setting, 0.056 for the urban slum setting, and 0.069 for the urban mixed/non-slum setting. A comparison, on the one hand, with the estimates of *Q* for other LMICs countries (e.g., 0.051 in Zimbabwe^7^, and 0.075 in Vietnam^3^ and Peru^2^, own calculations), and, on the other hand, with those from high-income countries in Europe (ranging from 0.14 in the United Kingdom to 0.19 in the Netherlands ^34,35^) clearly showed the higher degree of mixing between different age groups (and the lower assortativeness) in Kenya and in other LMICs compared to high-income countries. Although contacts with children (age group 0-14) were reported by participants of all ages, their intensity was higher among child participants. Higher contacts with children across all age groups were especially reported in the rural setting (Figure 2C), possibly because of the higher rate of intergenerational residence and the larger HH size (Supplementary Fig. S3). A strong mixing between adults older than 20 years and younger than 60 years was also found across all settings.

**Figure 1.**
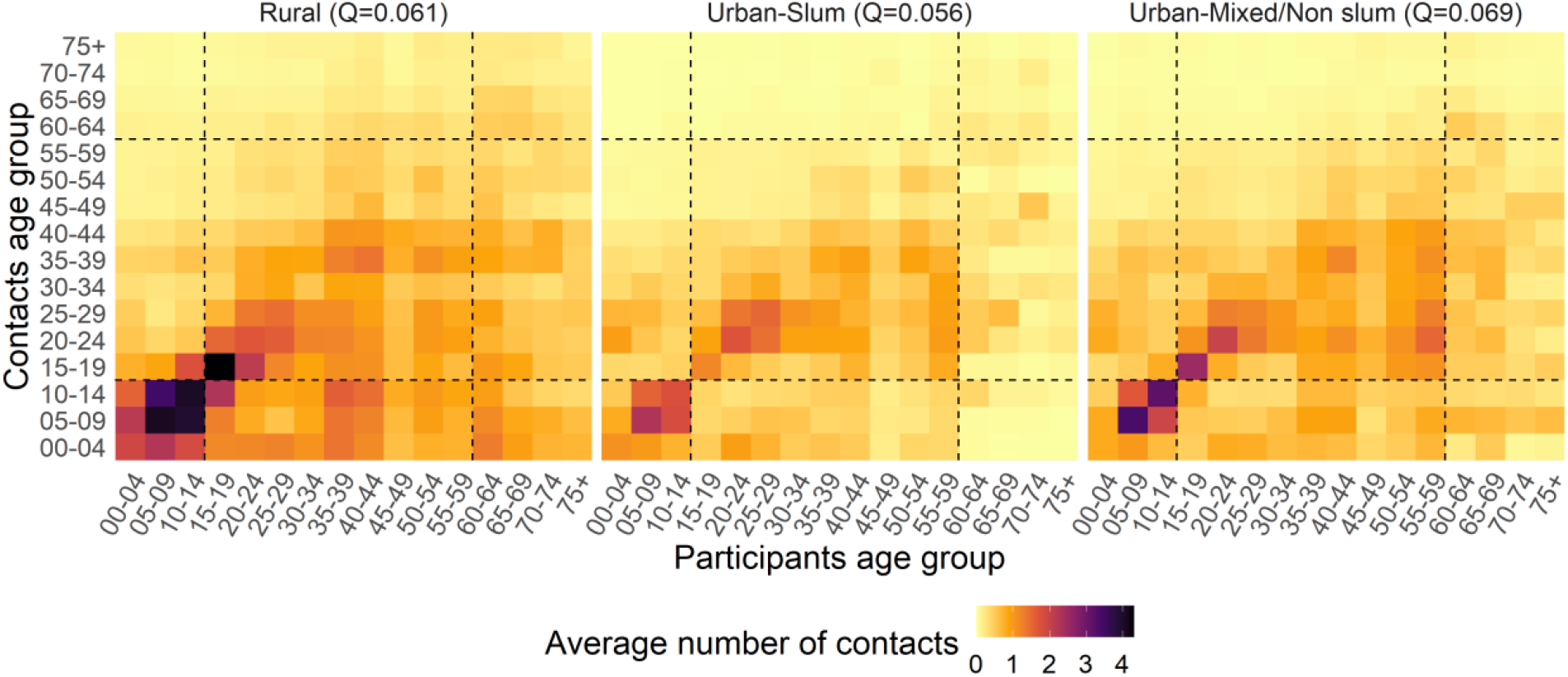
Average number of overall contacts between participants in age class *i* and contacted individuals in age class *j*, adjusting for reciprocity of contact at the population level, in each of the three settings. The dashed lines help identifying the contacts of children (0-14 years), teens and adults (15-59 years) and older adults (60+ years). At the top of each matrix, we report its assortativity index *Q*.

**Figure 2.**
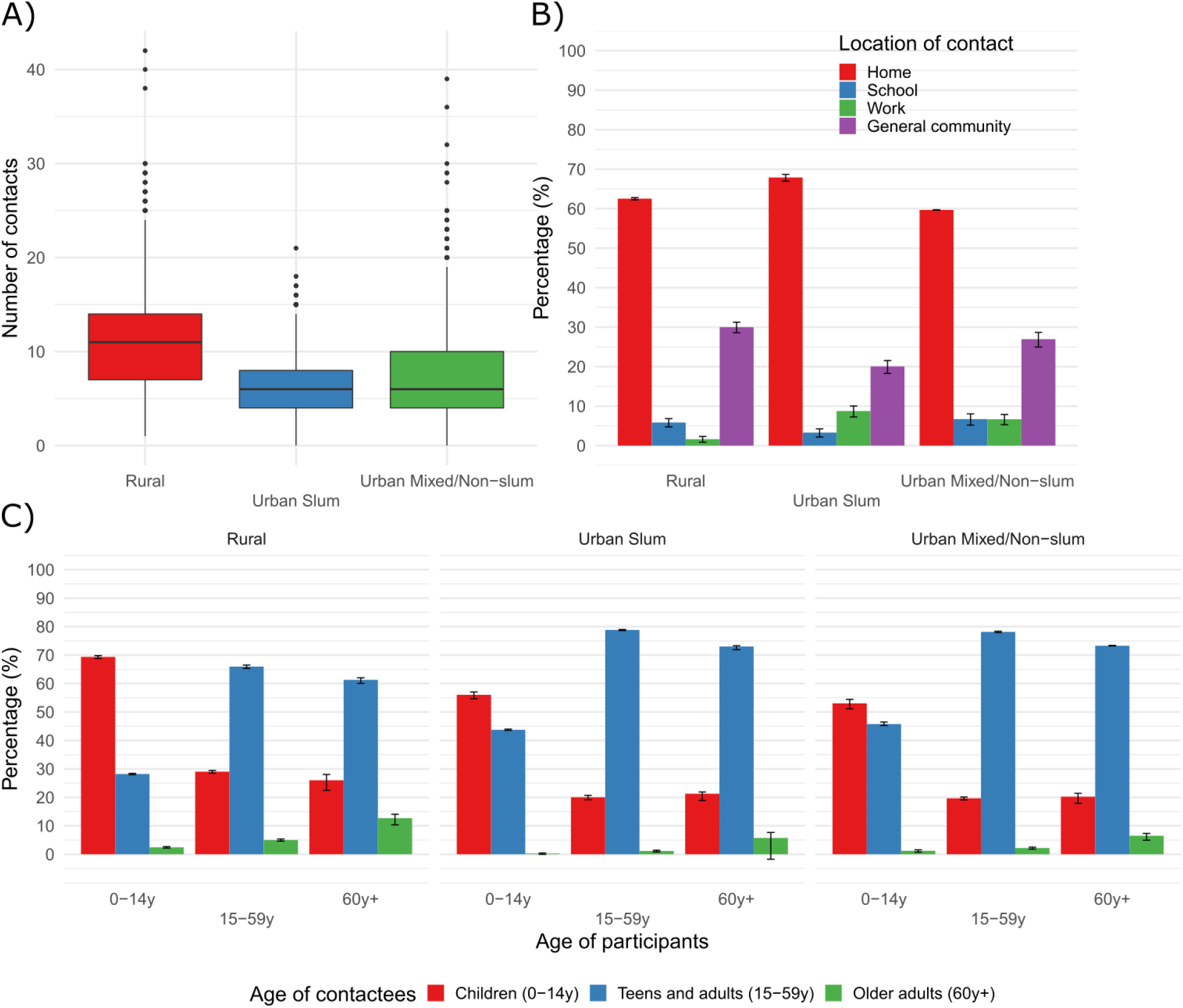
Distribution of daily social contacts by setting (A), percentage (with 95% CI) of daily social contacts by location of contact (B), and percentage (with 95% CI) of daily social contacts by setting and age of participants with children (0-14 years), teens and adults (15-59 years) and older adults (60+ years) (C).

Overall, the contact distribution was right-skewed, especially in the rural setting, where individuals in the last decile of the distribution reported between 18 and 42 contacts per day, while only between 12 and 21 in the slum (Fig. 2A). Regardless of the setting, most contacts were reported at home (from 59.7% in the mixed area to 67.9% in the slum) and in the general community (between 20.1% in the slum and 30% in the rural area) (Fig. 2B). Indeed, contacts at home and in the general community together represented 92.5% of all contacts in the rural setting, a share that was significantly larger than in both urban settings (87.9% in the slum and 86.6% in the mixed area), even after adjusting for participants’ characteristics (Supplementary Fig. S4).

Finally, across the settings, intergenerational mixing, defined as the mixing between different generations – children (aged 0-14 years), teens and adults (aged 15-59 years), and older adults (aged 60 years or more) – accounted for a higher share of total contacts of the adults aged 60 years or more (90.3%, of which 23.3%, CI 21.5%-24.6% with children) than for children younger than 15 years (37.1%, of which 1.7%, CI 1.5%-1.9% with older adults) or for teens and adults (26.9%, of which 3.2%, CI 2.9%-3.4% with older adults). Moreover, we found that the share of intergenerational contact reported with older adults was higher in the rural setting for both children (2.4%, CI 2.2%-2.7%) and teens and adults (5%, CI 4.6%-5.4%) (Fig. 2C).

### Differences in participants’ daily routines

To understand the behavioural determinants of the number of social contacts, six profiles based on individuals’ daily routine behaviour were identified by, first, applying principal component analysis (PCA) to the collected time-use (TU) data and, next, by applying a hierarchical cluster analysis to the retrieved principal components to group individuals into meaningful profiles ^36^. Each generated TU profile group can be described based on the most frequently visited location during the day, in terms of time slots, by individuals included therein (Fig. 3).

**Figure 3.**
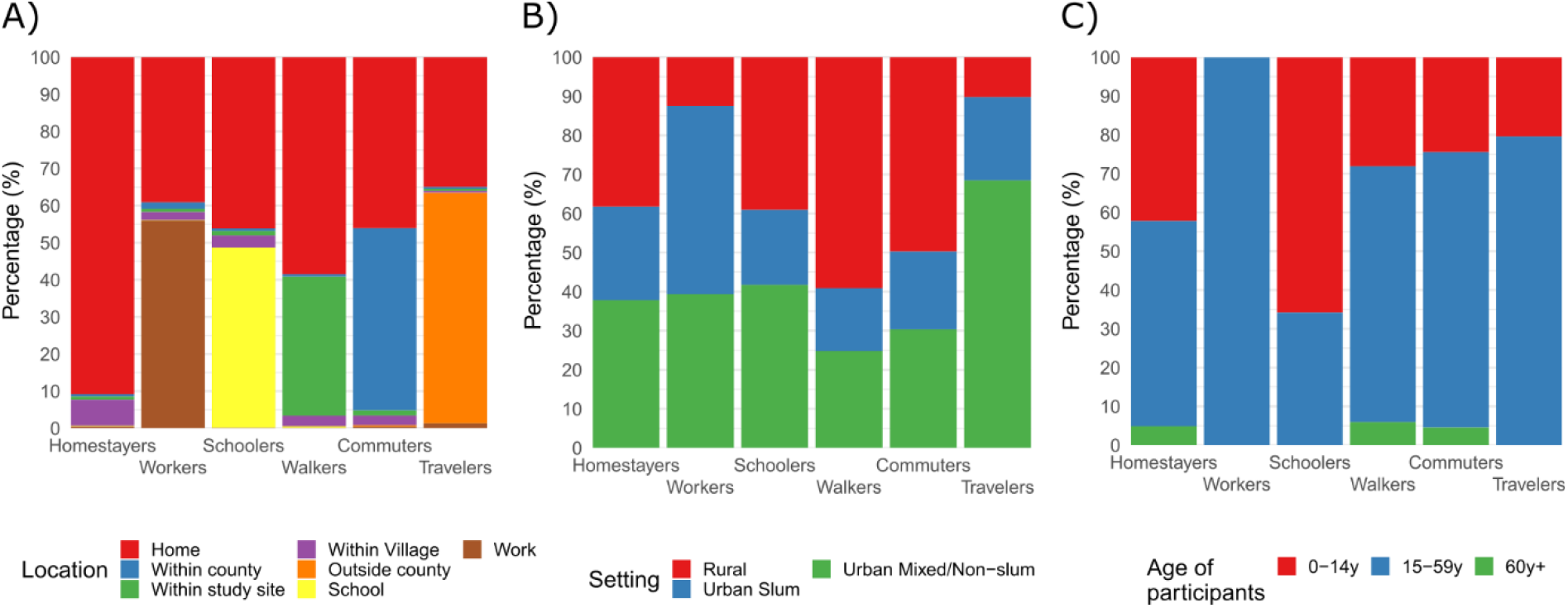
Description of the six TU profile groups by location (A), setting (B), and age group (C).

Most individuals (92.3%) reported the same TU profile over the two survey days, especially in the rural site (99%). The largest group by far (*Homestayers*) accounted for 71.1% of p.d. and identified people *staying at home or within the village* the whole day (respectively 90.8% and 7% of their daily time). The remaining TU profiles were characterized by a specific location, other than the home. The second largest group (*Workers*: 9.3% of total p.d.) identified those spending over half of their day *at work* (56%); this profile was more prevalent in the urban settings, especially the slum, and only for the group of adults. The third profile group (*Schoolers*: 7.8% of p.d.) gathered those spending a large share of the day *at school* (48.6%), which was uniformly present in all settings, and mostly populated by children.

The last three groups encompassed individuals spending a relatively larger share of their time in the general community, and were gradually more prevalent in the urban settings and among teens and adults as the distance from home increased: the fourth group (*Walkers*: 6.6% of total p.d.) included those spending a good share of their time *within the study site* (37.5%); the fifth group (*Commuters*: 4.6% of the total p.d.) encompassed individuals spending a relevant part of their day *within their county* (49.1%); finally, the last group (*Travelers*: 0.6% of the total p.d.) identified individuals spending a relevant part of their day *outside their county* (62.2%).

### Social contact patterns by TU profile

The TU profiles described above were used to assess differences in contact patterns between people characterized by different daily routines.

With regards to age mixing, we found that the degree of age-assortativeness varied across TU profiles (Fig. 4). This was lower for Workers (*Q*=0.0030), Travelers (*Q*=0.023), and Commuters (*Q*=0.046), for which age mixing, apart from contacts between parents and their children, was scattered among teens and adults of different age. The *Q* index increased to 0.057 for Homestayers and 0.066 for Walkers, reflecting a moderate level of age-assortative mixing (therefore, suggesting marked intergenerational mixing) among children and adults up to the age of 30 years. Finally, the highest level of age-assortativeness (and thus the lowest level of intergenerational mixing) was found among Schoolers (*Q*=0.10), where the intensity of contact among children reached the peak.

**Figure 1.**
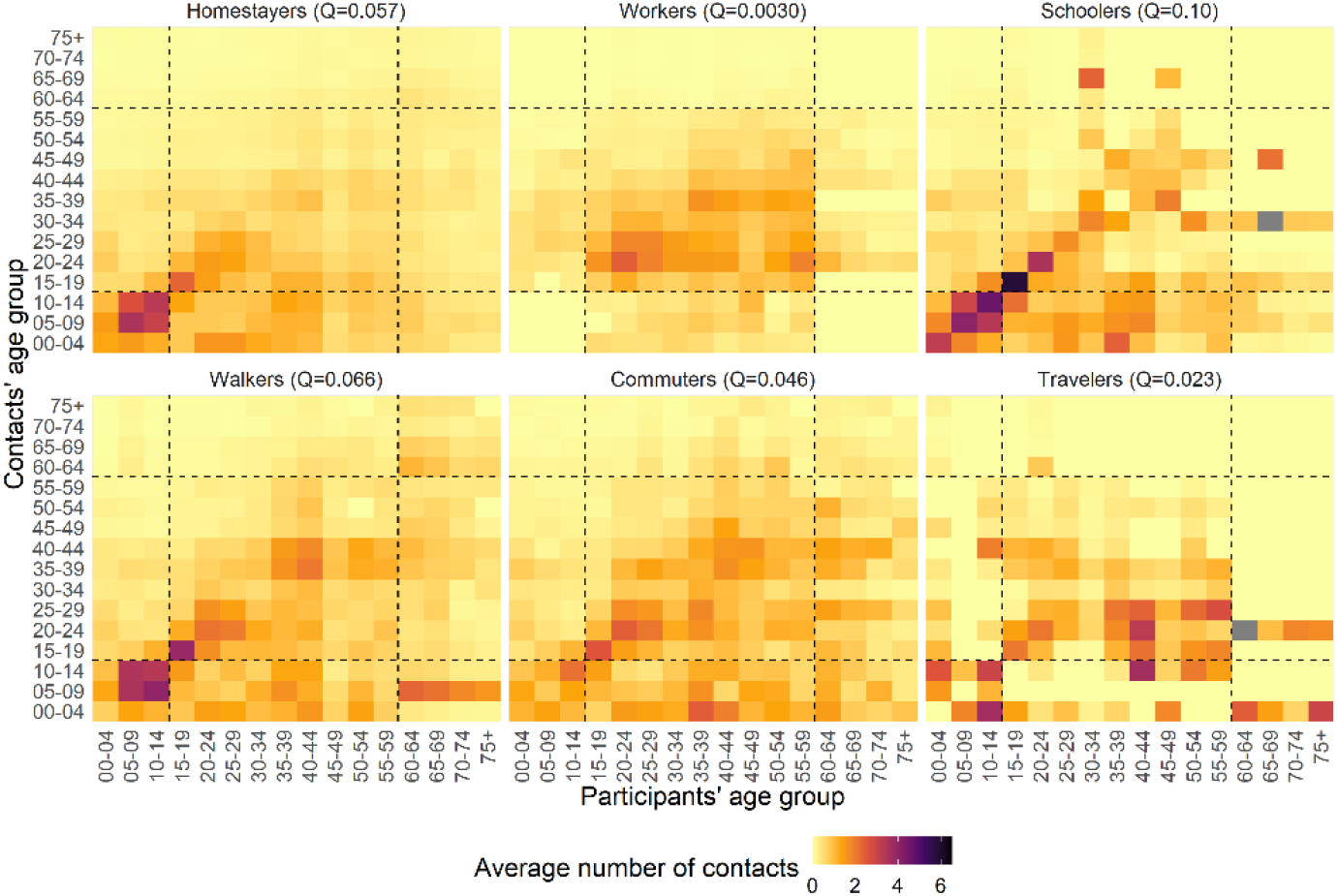
Average number of overall contacts between participants in age class *i* and contacted individuals in age class *j*, adjusting for reciprocity of contact at the population level, stratified by TU profile. The dashed lines help identifying the contacts of children (0-14 years), teens and adults (15-59 years) and older adults (60+ years). At the top of each matrix, we report its assortativity index *Q*.

The contact distribution under each TU profile was generally right-skewed, especially for Schoolers, who also reported a higher median number of contacts (Fig. 5A). Workers reported 43.3% of their contacts at work, Schoolers 48.5% of contacts at school, while Walkers, Commuters and Travelers reported between 55% and 64.4% of contacts in the general community, showing a positive relationship between the time spent and the reported number of contacts in each location (Fig. 5B). Contacts at home usually represented the largest share, except for Commuters and Travelers, who spent a larger share of time in the general community at increasing distance from home (Fig. 3A, 5B) and reporting a higher number of contacts as occurring in this location.

TU profiles also varied in terms of intergenerational mixing (Fig. 5C). Contacts with children were mainly reported by Schoolers (55.4%), followed by Homestayers (43.2%) and Walkers (34%). Within these three profiles, contact with children was mainly reported by child participants; however, we also observed a high share of intergenerational contact reported by older adults with a Homestayer profile (26%). Conversely, contacts of Workers with children did not reach 15%. Contacts with older adults represented a small fraction for all TU profiles, although we found significant differences between Workers and Schoolers (less than 2% of contacts), Homestayers (almost 3% of contacts), and Walkers and Commuters (almost 5% of contacts). Such contacts with older adults were mainly reported by participants of the same generation; however, we also observed a substantial level of intergenerational contact reported by both children (at least 3.2%) and teens and adults (between 4% and 5%), suggesting that interactions with the older adults occurred mostly with those individuals spending a high proportion of their time in the general community.

After adjusting for participants ‘ characteristics, we estimated that Workers and Schoolers reported overall 38% and 24% more contacts than Homestayers, respectively (Table 1). We did not find any significant difference in contacts between Workers and Homestayers at home and in the general community (Supplementary Fig. S6). Conversely, Schoolers reported less contacts than Homestayers both at home and in the general community, supporting the finding that most of their contacts occurred at school. Finally, Walkers and Commuters (who reported 10% and 21% more contacts than Homestayers, respectively) had less contacts at home and much more in the general community.

**Figure 2.**
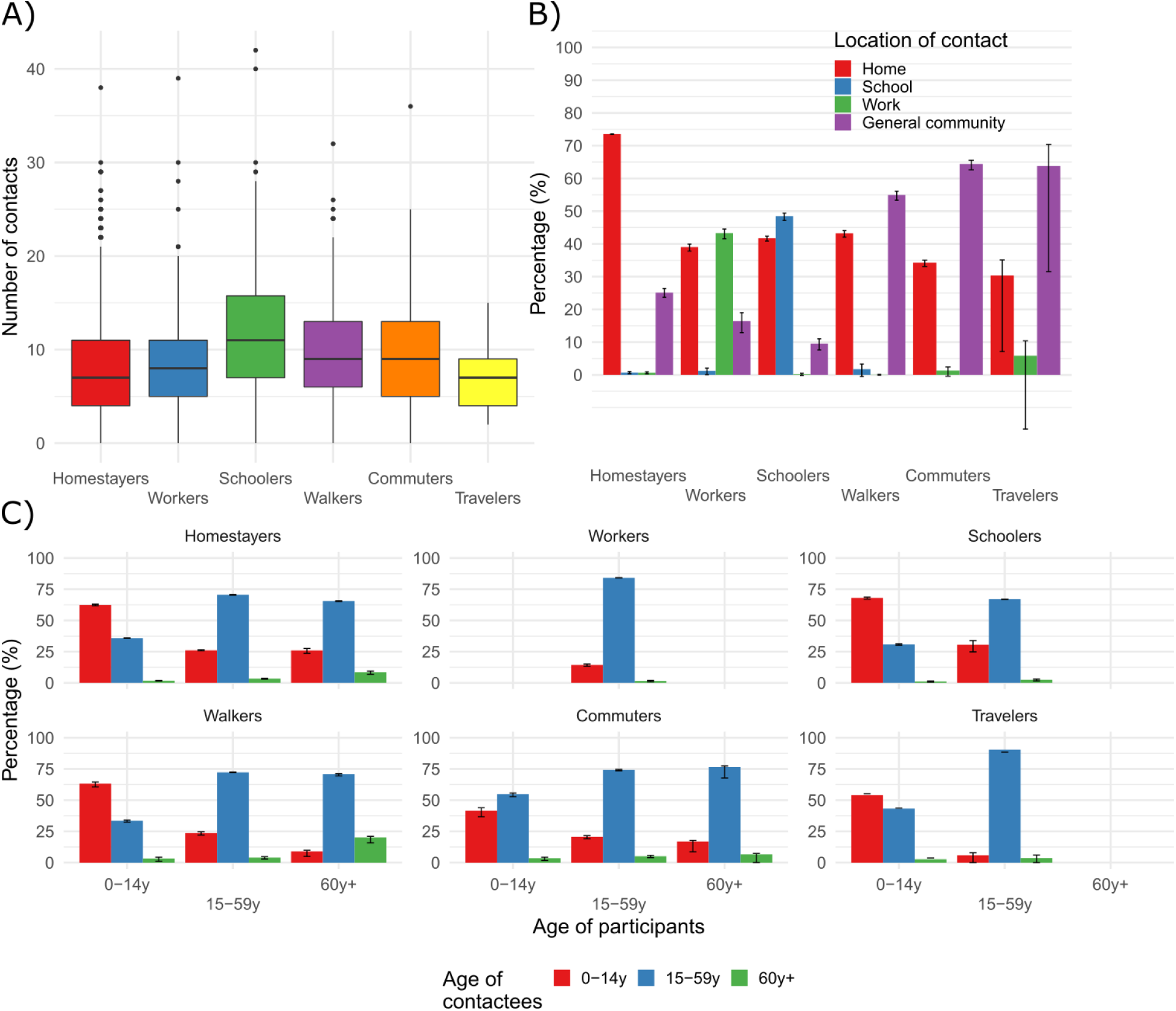
Distribution of daily social contacts (A), percentage (with 95% CI) of daily social contacts by location of contact (B), and percentage (with 95% CI) of daily social contacts by setting and age of participants with children (0-14 years), teens and adults (15-59 years) and older adults (60+ years) (C).

## Discussion

Although most social contact studies in LMICs ^2,3,6^ have been predominantly conducted in rural settings, while those conducted in high-income countries^31,34,35,37–42^ have been largely carried out in urban settings, only a few have so far presented a direct comparison of social mixing patterns between such different levels of urbanization within the same country or region ^1,5,7,8^. Moreover, only one study sought to investigate whether differences in social behaviour induced by daily routines entailed differences in the average numbers of contacts^7^. Our contact study in Kenya contributes to filling this gap by (i) providing a robust comparison of social mixing patterns across diverse demographic settings along the rural-urban gradient, and (ii) identifying which individuals ‘ profiles, based on their age mixing patterns and daily time use behaviour, can play a pivotal role in bringing different age groups in contact. The latter aspect is of great importance to identify those individuals who present a higher risk of bringing an infection to the more vulnerable segments of the population.

Even though this study was not the first one conducted in Kenya^5,43,44^, it was the largest and most diverse one in terms of surveyed settings, describing three geographical areas that significantly differed in terms of socio-economic status (the rural setting being the poorer in relative terms, the urban mixed/non-slum area being the wealthier), age and household size distribution (a younger population and a higher frequency of large extended households in the rural setting, and a higher frequency of small nuclear households in the slum), and TU profiles (a higher percentage of people spending time at work in the urban settings, especially in the slum, as well as a higher percentage of people spending time in the general community in the rural setting).

The number of contacts reported by our study participants were higher in the rural setting than in the urban ones, even after adjusting for the HH size (higher in rural site) and the living arrangements (with higher intergenerational cohabitation in the rural site), as a result of the higher contact numbers reported at home and in the general community. Even though these findings differed from what was found for Zimbabwe ^7^, they were consistent with what was observed by previous studies in the same area in Kenya ^5^, in Zambia and South Africa ^1^, as well as with the evidence from the synthetic matrices for low-income countries constructed using setting-specific survey data (e.g., on household, school, classroom, and workplace composition, and on contact patterns) ^45^. Despite the lower degree of age assortativeness in comparison to high-income countries, regardless of the type of setting, we observed both a higher intensity of contacts among children and a higher share of intergenerational contact with older adults in the rural setting than in the urban ones (Fig. 2C). Such mixing behaviours should be taken into account when considering the transmission of infections with strongly age-specific severity, such as in the case of RSV or SARS-CoV-2^21,22,26,28^.

Compared with previous studies conducted in Kenya, we found that our survey participants reported a lower number of contacts: a study carried out between 2011 and 2012 in Kilifi county reported 18.8 physical contacts per day in a rural setting and 16.5 in a semi urban area ^5^, while a small study conducted in a slum in the capital Nairobi in May 2020 found an average of 18 overall contacts per day^44^. Such discrepancies with our study might be due to differences in the sample selection, especially in the urban site. First, since we lacked a sampling list for our urban site, conversely to what happened with the study in the Nairobi ^44^ slums, we employed a modified version of the WHO Expanded Program on Immunization (EPI) cluster sampling technique ^46^ (see supplementary materials), which may have caused some bias in the household selection. Second, the inaccessibility to some homes, especially the guarded houses in the non-slum area, may have been another possible source of bias in the household selection. Finally, the requirement that interviewers worked between 7am and 6pm, due to both security measures and working time restrictions, may have prevented the inclusion of working adults who might be outside the home when interviews were conducted. On the other hand, to improve data quality, we used face-to-face interviews conducted by a trained interviewer instead of self-reporting paper diaries ^5^ or phone interviews^44^ to improve data quality, especially for those collected from illiterate respondents.

The combined collection of time-use and contact data within the same study allowed us to investigate how people differed in terms of daily behaviour and how their behaviour was associated with a distinct age mixing and contact numbers. At least 70% of participants spent most of their day at home or in the village, without any time at work or at school, and reported the lowest number of contacts (8.45 on average), 74% of which on the premises of their home. Intergenerational mixing patterns were found to be less relevant for Workers and Schoolers and more substantial for Homestayers, Walkers and Commuters. On the one hand, the level of intergenerational mixing increased for people spending more time farther from home, suggesting that people who spend most of their time and report most of their contacts in the general community might be relevant for the transmission of COVID-19 as they are characterized by a larger number of interactions between teens and adults and with older adults. These behaviours may be those mostly affected by social distancing and isolation, which are strategies designed to reduce the number of contacts with the most vulnerable age groups. On the other hand, the rather large mixing reported by older adults with children at home may be more difficult to control, as people who spend most of their time at home are less likely affected by public restrictions targeting individual movement, social activities, and school closures.

In conclusion, our results highlight that the higher number of reported contacts and the higher level of intergenerational mixing, especially between children and older adults in the rural site, as opposed to the urban slum, and mainly reported by individuals who spend most of their day at home or in the general community, may have important consequences for COVID-19 disease transmission to older adults. More in general, considering the heterogeneity in contact numbers and social mixing entailed by differences in time use behaviour, rather than simply looking at the variations in contact patterns per location, may achieve a better identification of groups of individuals at higher risk of transmission, define effective measures to prevent the spread of the diseases, and design appropriate targets of future vaccination efforts.

## Materials and Methods

### Study population

The rural site was selected among the most rural villages within the Kilifi Health and Demographic Surveillance System (KHDSS), an area including most of patients admitted to Kilifi District Hospital. The HDSS had an average population of 261,919 between 2006 and 2010, a population growth rate of 2.79% per year, and a total fertility rate (TFR) of 4.7 ^29^.

On the other hand, the urban site of Mombasa County, had a population of 939,370 in 2009 and in 2014 reported a population growth rate of 3.60% per year ^47^ and a TFR of 3.2^48^. Mombasa County was also characterized by disparities in living conditions within the city and by the presence of informal settlements (slums), which featured higher fertility and mortality compared to the rest of the country^47^.

### Study design

The study design was cross-sectional, targeting recruitment at the household level. Individuals of all ages were grouped into seven age classes reflective of key social or behavioural groups: <1 year (infants), 1–5 years (preschoolers), 6–14 years (primary school), 15–18 years (secondary school), 19– 34 years (younger adults), 35–59 years (middle-aged adults), and >60 years (older adults).

Estimates from a previous contact study conducted in Kenya were used to parameterize the sample size calculation ^5^. To account for the larger heterogeneity in the social structure of the urban area in terms of household composition and social interactions, 1000 participants were targeted in the urban area and 500 from the rural area. Although the sample size was allocated to the seven age groups proportional to the population of the corresponding age strata, children were oversampled due to the critical role they were expected to play in infectious disease transmission and hence the need to reduce the level of uncertainty in the estimated contact rate for this age group. Recruitment was staggered over a six-month period (June 2015 to December 2015). Inclusion criteria included (i) giving informed consent, either directly or by parents if participants were minor of age, and (ii) planning to remain in the site for at least two weeks. Information sheets and consent forms were provided to all participants.

Background information on the participants regarded their age and sex, their education career and occupational status, their HH size and characteristics (such as owned assets, use to derive a SES measure), and their school and work environments (e.g., distance from home, school class and workplace size).

Similarly to previous studies ^7,34^, contacts were defined as an interaction between two individuals, either physical (involving skin-to-skin contact), or non-physical (involving a two-way conversation with three or more words in the physical presence of another person, but no skin-to-skin contact). For each contacted individual, participants provided information on the sex and the age of the contacted person, the type of contact, the location(s) where the contact occurred (home, school, work, general community), and the relationship with the contacted person (e.g., sibling, parent, non-relative). To distinguish short-lived contacts with long-duration contacts, and to account for the possibility that an individual may be encountered several times during the day, we collected the information on the total duration of the contact with such an individual.

Finally, participants provided information on their time-use by recording all the visited locations (the same used for contacts) during their day. For this purpose, each survey day was divided into nine time slots reflecting the position of the sun, spanning from a minimum of two hours (e.g., dawn, late morning, or dusk/early night) to a maximum of eight hours (late night). Respondents reported whether, for each time slot, they had visited or not a given location (home, school, work, and general community – within the village/estate, within the study site, within the county, or outside the county).

The study was approved by the Kenya Medical Research Institute-Scientific Ethics Review Unit (KEMRI-SERU) and the University of Warwick Biomedical and Scientific Research Ethics Committee (BSREC). The parent study, DECIDE, had received ethical approval by the ERC Ethics Committee and the Ethics Committee of the Italian National Institute of Health. Individual consent was obtained from each of the study participants.

### Statistical analysis

We analysed all collected data by adjusting for sampling weights (the inverse of the probability that an observation is included because of the sampling design), which were calculated for each site separately to compute site-specific estimates. These weights were based on both the KHDSS and the Mombasa County population age distribution, taken as reference populations.

We created two new variables for inclusion in the statistical models: a continuous SES index and a TU profile variable. The SES index was computed by applying PCA to data on household characteristics and owned assets, also collected in the study, and taking the first component as the index^49^. To construct the TU profiles, we applied PCA to the collected TU data to summarize all the information contained in the variables with a smaller number of dimensions, constructed to be independent of each other. We used the obtained components as input for a hierarchical cluster analysis to create groups of person-days characterized by similar time use behavior profiles^36^.

Social contact matrices by study site and TU profile were constructed to show the average number of contacts between 5-year age classes in the respective populations (plus a single age class from 75 years or more). Each matrix element, 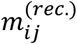, contained the mean number of contacts per day in age class *j* as reported by participants in age class *i*. To take into account the reciprocal nature of contacts^50^, the elements of the matrix were adjusted for reciprocity at the population level (see the supplementary materials for further details on the construction of the contact matrices).

We measured the age-assortativeness with the *Q* index^31,33,51^, which is calculated as *Q* = [*Tr*(*P*) − 1]/(*n* − 1), where *P* is the matrix whose elements represent the fraction of total contacts of age group *i* with age group *j, P*_*ij*_ = *M*_*ij*_/*∑*_*j*_ *M*_*ij*_, *M*_*ij*_ is the matrix with the number of total contacts between age groups, and *Tr*(·) is the trace of the matrix, i.e., the sum of its diagonal elements. The *Q* index takes values one as the assortativeness becomes maximal (all contacts are on the main diagonal, i.e., with individuals of the same age), while, for increasing homogenous (or proportionate) mixing, it tends to zero^32^.

Generalized estimating equations (GEE) were chosen to evaluate the effect of the different participants’ characteristics on the daily number of contacts ^52^. The negative binomial distribution was preferred over the Poisson to allow for a higher variance in the number of contacts reported by participants than what expected under the former distribution (overdispersion) ^7,34,53^. We tested, however, for the possibility of reducing the distribution to a Poisson by checking whether the 95% confidence interval (CI) of the estimated overdispersion parameter contained zero. Contact data were clustered within participants over the survey days under the assumption that responses from the same individual were correlated, while responses from different participants were assumed to be independent.

We fitted the GEE model to all contacts and then separately for each of the three settings. Except for the setting, which was included only in the overall model, the variables included in all models were the sex, the age group (0-14 years, 15-59 years, and 60 years or more), the HH size (approximately grouped in five quantile groups, i.e., 1-3, 4, 5, 6-7, and 7 or more individuals), the living arrangements (one-generation, two-generation, or three-generations HHs), the SES index, the current education level, the TU profile, and the type of survey day (weekday or weekend). We interpreted the exponentiated model estimates from the negative binomial GEE as incidence rate ratios (IRR), giving the relative number of contacts per day with respect to the reference category of each variable, and we accompanied them with 95% CI.

Data cleaning and wrangling, matrix construction, and result visualization were carried out in R (using, in particular, the package “FactoMineR” for the PCA and the HCA^54^); GEE models were estimated in Stata, using the procedure “xtgee”.

## Data Availability

The datasets and the source codes necessary for the reproducibility of our results are available at the following repository.

https://osf.io/xtw8h/?view_only=27b7ccf00a4247cea0a50b69617a988f

## Acknowledgements

We thank all the study participants for their contribution of time and data. We also thank the Tudor location community health volunteers and local administration in Mombasa for providing security throughout the data collection period. We are grateful to the field study team lead by Grace Jumbale and John Koi who dedicated their time to make sure data collection was successful. We acknowledge Edward Mundia, Lilian Mwango, and Grieven Otieno for the digital system development, the data clerks for data entry and management, and the Kilifi HDSS registry. This article is published with the permission of the Director of KEMRI. This work has received funding from the European Research Council under the European Union’s Seventh Framework Program (FP7/2007-2013)/ERC Grant Agreement 283955 and has been partially supported by the Wellcome Trust (Grant # 203077/Z/16/Z).

## Author contributions

**Emanuele Del Fava:** Conceptualization, Formal analysis, Investigation, Methodology, Software, Visualization, Writing – original draft, review & editing. **Irene Adema:** Data curation, Investigation, Resources, Writing – original draft, review & editing. **Moses C. Kiti:** Data curation, Investigation, Project administration, Writing – review & editing. **Piero Poletti:** Conceptualization, Investigation, Methodology, Writing – review & editing. **Stefano Merler:** Supervision, Writing – review & editing. **D. James Nokes:** Funding acquisition, Supervision, Writing – review & editing. **Piero Manfredi:** Conceptualization, Supervision, Writing – review & editing. **Alessia Melegaro:** Conceptualization, Funding acquisition, Project administration, Supervision, Writing – review & editing.

## Data availability

The datasets and the source codes necessary for the reproducibility of our results are available at the following [blind for peer-review] repository: https://osf.io/xtw8h/?view_only=27b7ccf00a4247cea0a50b69617a988f.

