## Supplementary for "Individual’s daily behaviour and intergenerational mixing in different social contexts of Kenya"

#### Sampling Strategy

##### Study area

Two sites were selected, one each from a rural and urban setting at the Kenyan coast. The rural site was located within the Kilifi Health and Demographic Surveillance System (KHDSS)[1], while the urban site was within Mombasa city in coastal Kenya. The rural-urban population density ratio was 1:56 (2009 national census data), suggesting very high population density in the urban area.

The rural area was characterized by large families with a high proportion of children and high out-migration rates in adults in search for work, mainly in urban areas. Residents lived in homesteads, which are groups of related households living within the same compound. The main economic activity is subsistence farming with cyclic patterns following seasonal rainfall patterns experienced between March and May and between October and December.

On the contrary, the urban location had two distinct settlement patterns. The first was a slum area dominated by high-density informal settlement structures with nuclear families living in each household. The second was middle-class housing and estates nested within gated communities that were more difficult to access due to seclusion. The urban site was bordered by two major highways, making it an important transit point to and from Mombasa. There were also several primary and secondary schools, a public university, and business premises especially, hotels for tourists.

##### Study design

The study design was cross-sectional, targeting recruitment at the household level. The study was designed to span various time points, social and economic activities, such as school term and holiday periods, and seasonal farming activities that are expected to influence contact patterns. Individuals of all ages were expected to participate, with participants grouped into seven age groups reflective of key social or behavioural groups: <1yr (infants), 1–5yrs (pre-school), 6–14yrs (primary school), 15–18yrs (secondary school), 19–34yrs (younger adults), 35–59yrs (middle-aged adults) and >60yrs (older adults).

##### Sample size estimation

Estimates from a previous contact study conducted in Kenya were used to parameterize the sample size calculation equations[2]. We aimed for an overall sample size of 1500 individuals. To account for the larger heterogeneity in the social structure of the urban area in terms of household composition and social interactions, 1000 participants were targeted in the urban area and 500 from the rural area. With these two sample sizes, using a t-test of power 78%, we would be able to detect a difference of 15% between the average number of contacts of the two sites, having as an alternative hypothesis that these are different. Considering a refusal probability of 50% (conservative value), reported in the previous contact study in Kenya[2], recruitment was carried out continuously with replacement of individuals who declined to consent until we attained the set sample size in both sites. Sample size was allocated to the seven age groups proportional to the population of the corresponding age strata. Infants were oversampled due to the critical role they play in the burden of childhood infectious diseases and hence the need to reduce the level of uncertainty in the estimated contact rate for this age group.

**Table S1 Sample size allocation matrix.** Sample size by study site and age group (N), with power to detect a difference between the expected estimates from this study and those found in a previous study[2], and census/sample ratios (ratio of population size to sample size, if C/S <1, we have oversampling).

| Age class (years) | Banda ra Salama (rural) | | | Tudor, Mombasa (urban) | | |
|---|---|---|---|---|---|---|
| | N | Power | C/S | N | Power | C/S |
| <1 | 50 | 0.54 | 0.43 | 100 | 0.83 | 0.35 |
| 1-5 | 67 | 0.39 | 1.06 | 98 | 0.53 | 1.09 |
| 6-14 | 137 | 0.67 | 1.06 | 108 | 0.57 | 1.07 |
| 15-18 | 48 | 0.30 | 1.06 | 100 | 0.54 | 1.07 |
| 19-34 | 90 | 0.50 | 1.06 | 358 | 0.97 | 1.07 |
| 35-59 | 71 | 0.41 | 1.06 | 202 | 0.83 | 1.07 |
| 60+ | 37 | 0.24 | 1.06 | 34 | 0.22 | 1.03 |
| Total | 500 | | | 1000 | | |

#### Participant selection

The existence of a well-defined sampling frame in the rural site, the Kilifi Health and Demographic Surveillance (KHDSS) platform, enabled the use of stratified random sampling to select participants from the site. Participants were selected at random, proportional to age category, from the KHDSS population register.

A similar population register was not available for the urban site, hence a multi-stage sampling method was adopted, slightly modifying the WHO Expanded Programme on Immunization (EPI) cluster sampling technique[3]. The study site was divided into irregular grids based on community health units, with the help of a community health volunteer who was working with the local health center. Households were secondary sampling units and were selected using a modified “random walk” door-to-door recruitment strategy. The total number of participants selected from each grid was proportional to its population density estimated from satellite imagery.

Recruitment was staggered over a six-month period (June 2015 to December 2015). Inclusion criteria included (i) giving informed consent, either directly or by parents if participants were minor or age, and (ii) planning to remain in the site for at least two weeks. A contact was defined in two ways, either as a physical contact; involving a skin-to-skin interaction, such as a handshake, a hug or a kiss, or as a non-physical contact; involving a two-way conversation with three or more words in the physical presence of one another (standing at most at two-arm distance), but no skin-to-skin interaction[4].

#### Community engagement

Prior to data collection, the research team conducted a comprehensive risk assessment and developed a mitigation plan as part of the host institution procedures. This was done in response to travel advisories issued by the national government at the beginning of the study period due to security concerns at the coastal region of Kenya. Participants were expected to give sensitive personal details of whom they met, location of their interactions, and how they spent their time over two days. This necessitated intensive community engagement before starting the data collection. Sensitization meetings were held with the local administration, including the chiefs, the village elders, the community representatives, and the community members. At these community meetings chaired by the chiefs, discussions focused on the study recruitment plans, procedures and confidentiality of the data to be collected. To prevent risks such as theft of study property, assault, or harassment of study staff, especially in the urban slum, field staff residing in these locations and known to the local administration officers were recruited for data collection. A network of local community health volunteers was identified and trained to assist in the household recruitment process.

#### Study implementation

Participant identification and recruitment procedures differed in the two sites. Records of selected participants in the rural site were obtained from the population registers. Their global positioning system coordinates were extracted, and the houses mapped out. Field staff visited the houses and recruited participants at the household level.

The urban site was divided into grids according to administrative boundaries. Field staff selected central points within their allocated grids from a map. Each would then spin a pen to randomly select the initial direction for recruitment. One person from each fifth household based on the predetermined age stratum was invited to participate. They asked about individuals in the first age group and, if there was nobody in that age group in the house, they proceeded to the next age group. They would then revisit the age group that was missed in the next house selected. This would go on until the entire area was visited and all age categories filled.

Trained field workers explained the details of the study to potential participants. Guardians gave consent for all children below 18 years. Guardians were requested to respond on behalf of all participants under 10 years old and other individuals who were unable to read and write (established by asking literacy status of individuals aged over 10 years). This process involved keeping track of the participants movements and noting down the contacts. The guardians who assisted with this task were termed as “shadows”. Guardians coordinated the process with schoolteachers for participants who spent most of their time in school.

Once consent was given, the fieldworkers recorded socio-demographic information about the participant (occupation, education, family composition, sleeping arrangements, i.e., sharing of bedroom or bed), recording the answers into an electronic questionnaire coded into a mini laptop. The participants selected the day in which they would keep the diary, and, consequently, the day of filling in the retrospective diary and the exit interview, by randomly picking a card from a set of seven cards that represent the weekdays.

Each contact was recorded only once in the diary during the selected day. The total duration of repeat encounters, and the different settings contacts occurred were summed up and recorded. Participants were expected to keep the diary for two consecutive days; a day defined as the period between first waking and going to bed for the night.

One day prior to the selected day, the fieldworker visited the participants to remind them to keep a mental note of the people they encounter on the day they selected. On the appointed day, for each different individual physically contacted, participants recorded the assumed age category of the person contacted in the diary against a unique identity code. Field workers visited the participant after the initial 24 hours at an agreed time to record the contacts in the electronic diary. If the participant was not available, the fieldworker would come back after the 48 hours to record the contacts for both days. If the participant were not available on the second day, the fieldworker would go back within the next 12 hours and drop the participant from the study if they could not be reached then. In addition to giving details of the people they contacted, participants also gave details of the settings in which they spent the 48 hours in a time use questionnaire. A clinical assessment questionnaire was also administered to inquire about the health status of the participant on the days of diary keeping.

#### Informed consent by subjects and/or community leaders

Information sheets and consent forms were provided to all participants. The information sheet contained a short introduction and purpose of the study, how the data was collected, the risks and benefits to participants. The study was explained to all participants using the information sheet. Children aged 13 – 17 years had to indicate willingness to participate through verbal assent. This was documented through an assent form signed by the field workers. However, individual written consent was sought from their parents.

#### Confidentiality

Each participant was assigned a unique study identification number (ID) that was used on the questionnaires and diaries. Interviews were done in private, unless the participants requested otherwise.

No names were used. Codes known only to the participant were used rather than actual names to indicate the contacted persons. Data were stored in secure password-controlled databases and no individual identities were used in any reports or publications resulting from the study.

##### Risks

There were no significant risks associated with the study. However, sufficient measures were put in place to minimize any potential risks. Unique study IDs and contact-person codes known only to the investigators and participants respectively were used. The reminder forms remained property of the participants. All study tools were locked in cabinets at KEMRI WTRP site, while awaiting entry into a password-protected database.

Inconveniences such as consuming participants' time during consenting and diary filling were expected to disrupt participants' normal routine. Study staff scheduled visits at participants' convenience to mitigate this and adhered strictly to appointments.

Because of the heightened insecurity concerns in the coastal regions by the members of the public, the study team put in place a comprehensive scheme to ensure safety of both study staff and participants. All study staff were provided with emblazoned t-shirts and work badges for identification. Staff were adequately trained on communication skills; to be polite to respondents, but also be able to perceive any risks (such as respondent hostility). Each staff member also had a card with important telephone numbers (the Security Officer, Health and Safety Manager at KEMRI-WTRP, the local area Chief/Village Elder, and the Police Station contacts). Staff were also expected to provide the official telephone number of the Study Coordinator to a respondent when asked.

#### Estimation of age-specific social contact matrices

Social contact matrices were constructed to report the average number of contacts between individuals in the age group $i$ with individuals in the age group $j$, $m_{ij}^{(rec.)}$, using a constraint for the reciprocity of contact when considering the total number of contacts at the population level.

##### Imputing missing age for contacted individuals

To deal with the missing information on the exact age of individuals contacted by participants, multiple imputation techniques were used[5]. For each contacted individual reported by participants without information on the exact age (but with information on the age class), we imputed the missing age by drawing it randomly with replacement from the given age class, proportionally to the age distribution of those individuals for which participants did provide the exact age. This imputation was replicated multiple times, generating 20 complete data sets. The final estimates were obtained by averaging out the output of the analysis of the complete data sets, while the related standard errors were calculated accounting for both within-replication and between-replication variability[5].

#### Constructing contact matrices

The calculation of the expected average number of contacts required to implement a constraint of reciprocity of contact at the population level[6]. This constraint implies that, in a closed population, the total number of contacts from age class $i$ to age class $j$ must be equal to the total number of contacts from age class $j$ to age class $i$. Hence, if we had $N_i$ individuals in age class $i$ and $N_j$ individuals in age class $j$, reciprocity would entail that $m_{ij}N_j = m_{ji}N_i$.

For this purpose, we started by dividing the total number of contacts, $M_{ij}$, for the number of participants in each age group, $n_i$, i.e., $m_{ij} = M_{ij}n_i$. We then constructed the matrix with the total number of contacts between age classes at the population level, by multiplying the expected mean number of contacts reported by participants in the age group $i$ with contactees in the age group $j$, $m_{ij}$, for the total number of individuals in the age group $i$ in the population, i.e., $C_{ij} = m_{ij}N_i$. At this point, we adjusted $C_{ij}$ for reciprocity by averaging the total number of contacts in one direction, $C_{ij}$, with those in the other direction, $C_{ji}$, weighting for the sample size by age group[7], namely, $C_{ij}^{(rec.)} = (C_{ij}n_i + C_{ji}n_j)/(n_i + n_j)$. We finally obtained the expected average number of contacts under reciprocity at the population level dividing again by the population $N_i$, i.e., $m_{ij}^{(rec.)} = C_{ij}^{(rec.)}N_i$.

### Additional figures and tables

#### Additional information on participants' characteristics

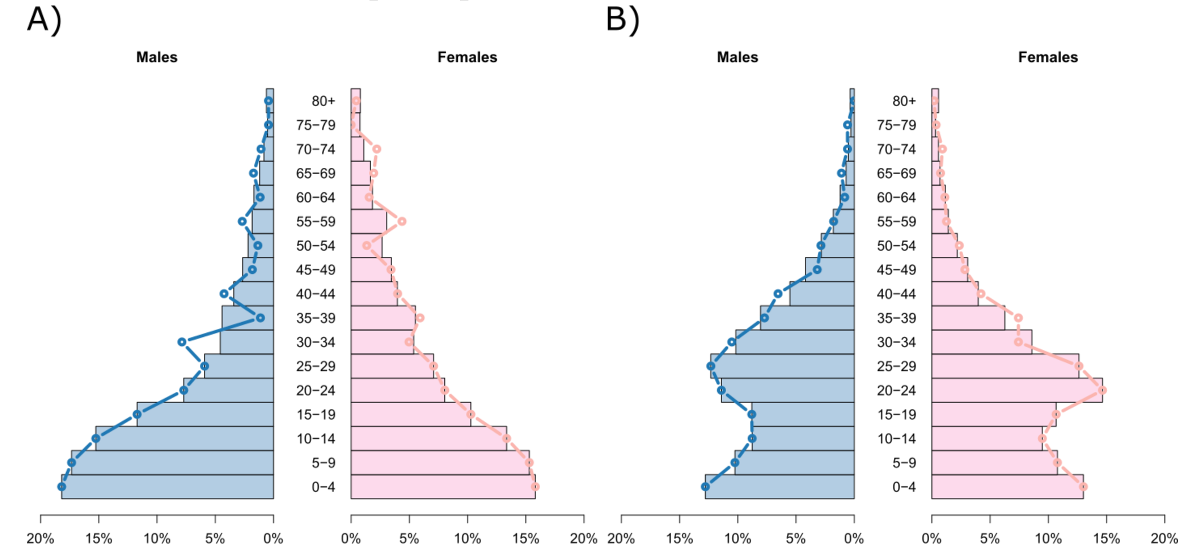


**Fig. S1 Age distribution by study site and gender.** Age pyramids of the population age distribution by gender (bar plots), overlapped by the weighted sample age distribution by gender (lines), in the rural site (A), and in the urban site (B), in 2015.

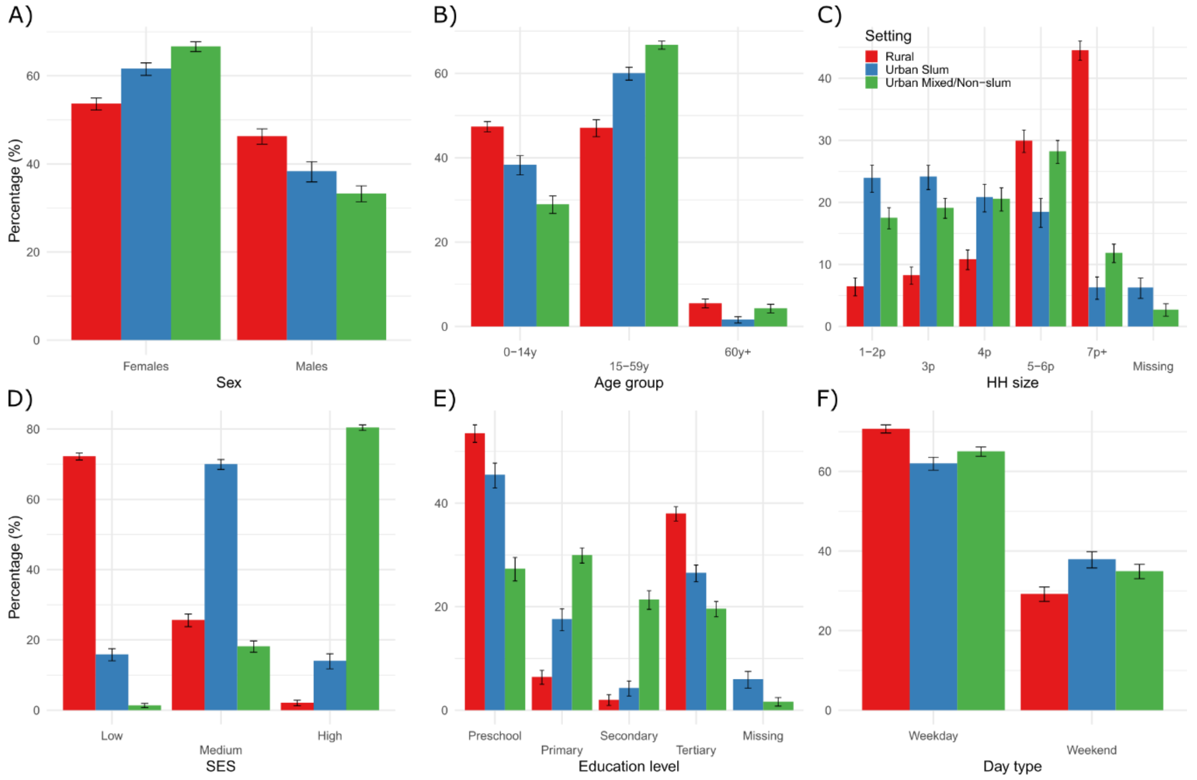


**Fig. S2 Participants' characteristics by setting.** Weighted percentage distribution of person-days by participant's sex (A), age group (B), HH size (C), SES index (D), current education level (E), and type of survey day (F), in each of the three settings.

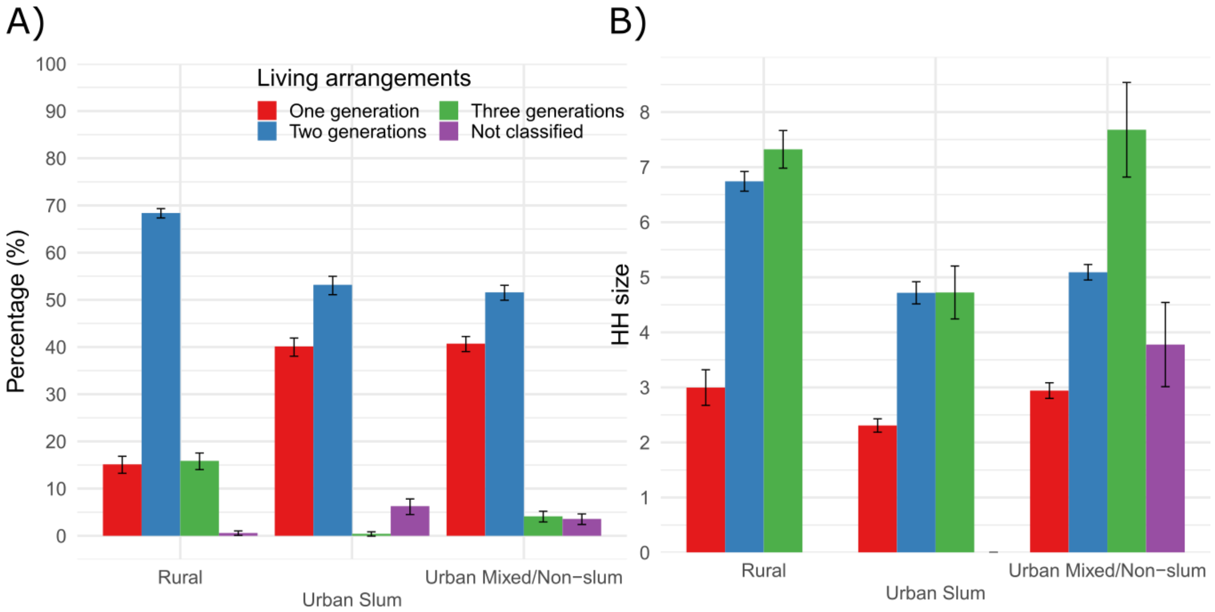


**Fig. S3 Living arrangements by setting.** Percentage of participants living in a home with people from different numbers of generations (children – 0-14y, teens and adults – 15-59y, older adults – 60y+) by setting (A); household size distribution by living arrangements and setting (B).

In terms of living arrangements, we found that intergenerational HHs, either with two or three generations living together, were more prevalent in the rural setting; on the other hand, one-generation HHs (i.e., couples living together without children) were more frequent in both urban settings (Fig. S3, panel A).

Moreover, the HH size increased with the number of generations living together across all settings. This finding was more evident in the urban mixed/non-slum area and, to a lesser extent, in the rural area; on the contrary, in the slum, we did not find any difference in HH size between the intergenerational HHs with two or three generations living together (Fig. S3, panel B).

#### Additional information on contacts' characteristics

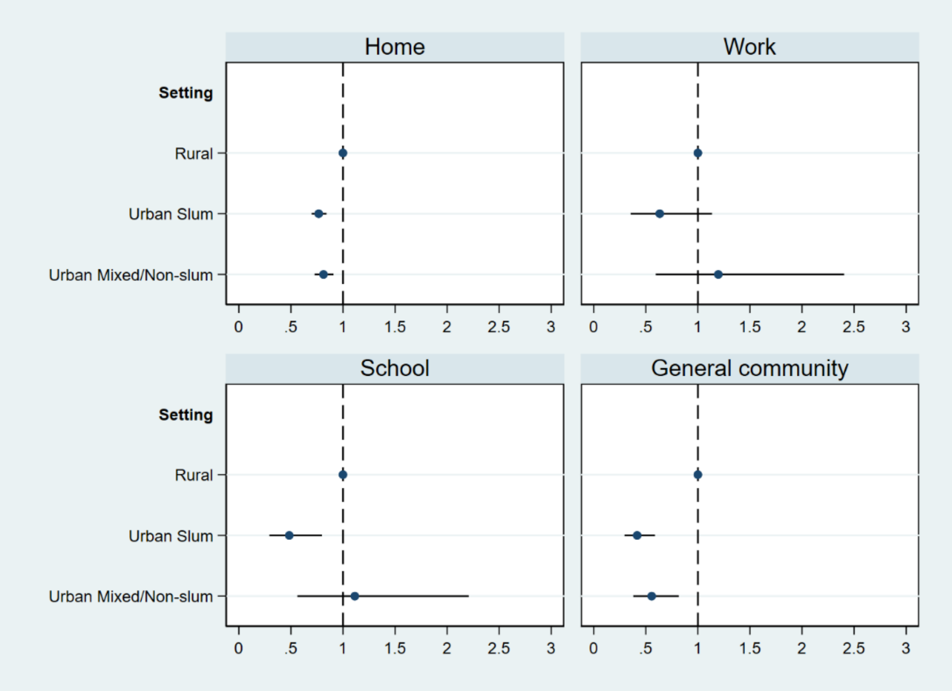


**Fig. S4 Association between setting and location of contact.** Relative number of reported contacts (IRR, with 95% CI) in each location, by setting of residence of participants. The estimates are obtained from a GEE with negative binomial distribution, adjusted for participants' characteristics (sex, age, education level, HH size, SES status, TU profile, and day type). The models for work and school contacts were run only within the subset of individuals with profile 2 (work) and profile 3 (school), respectively.

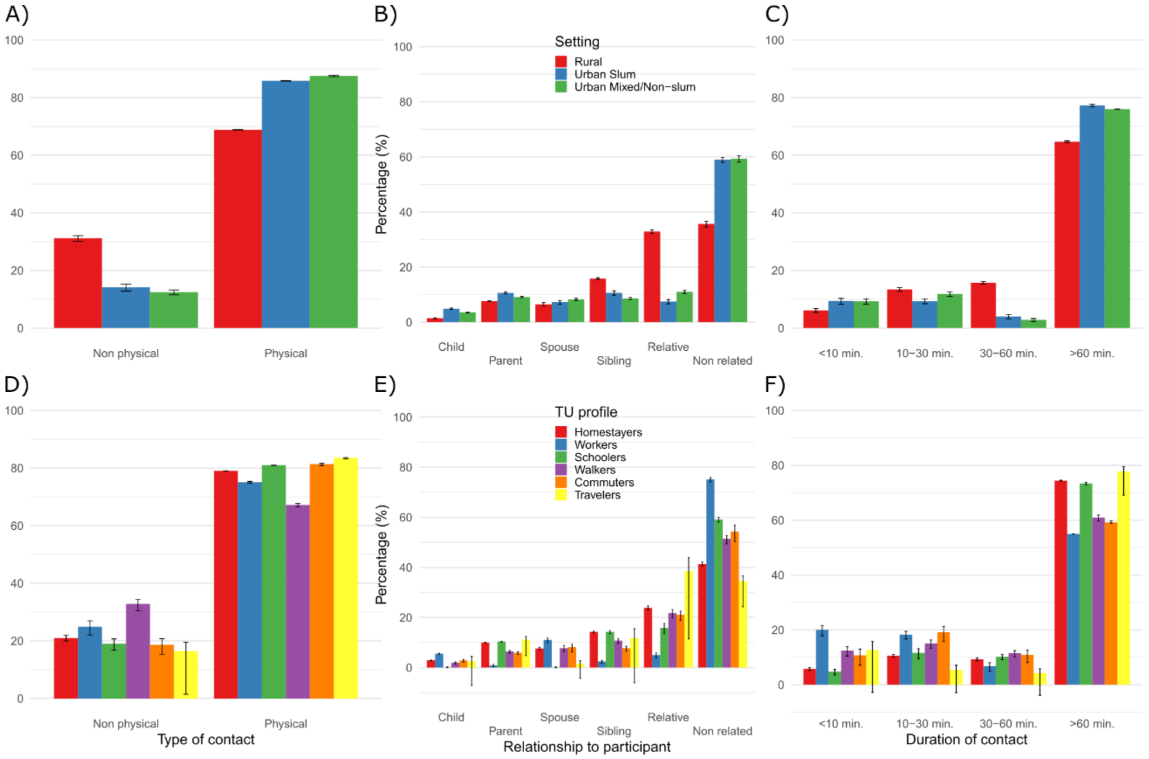

**Fig. S5 Characteristics of social contacts by setting and TU profile.** Weighted distribution of social contacts by relationship to the participant (A, D), type of contact (B, E), and duration of contact (C, F), in each of the three settings (A-C) and for each of the six TU profiles (D-F).

Most of the contacts were physical and of long duration. Regarding the setting, we found significant differences in the distribution of non-physical contacts (higher in the rural setting), of contacts with relatives (much higher in the rural setting), and of contacts with non-related individuals (much higher in both urban settings) (Fig. S5).

Considering the TU profile of participants (Fig. S5), Workers mainly reported contacts with not-related individuals – likely, colleagues and customers – in addition to those with children and spouse; Schoolers reported contacts with not-related individuals – likely school mates and teachers – as well as with parents, siblings, and other relative. Finally, Walkers, Commuters and Travelers in the general community reported large shares of contacts with not-related individuals with respect to Homestayers. In terms of duration, long duration contacts were mainly reported by Homestayers and Schoolers, while larger shares of contacts of short duration were primarily reported by Workers.

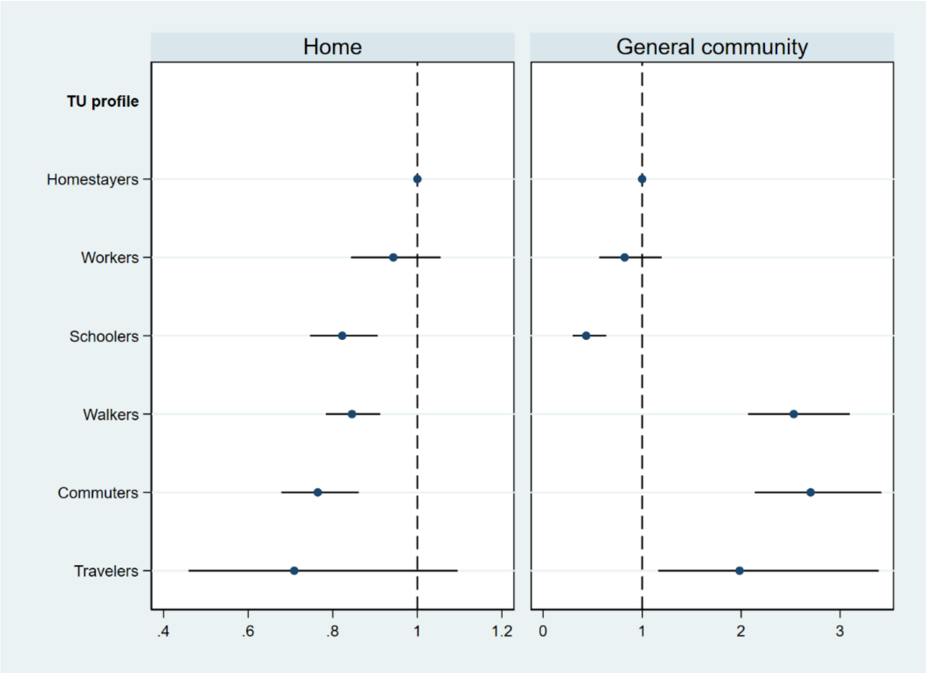


**Fig. S6 Association between TU profile and location of contact.** Relative number of reported contacts (IRR, with 95% CI) at home and in the general community, by TU profile of participants. The estimates are obtained from a GEE with negative binomial distribution, adjusted for participants' characteristics (sex, age, setting, education level, HH size, SES status, and day type). Results for the models for work and school contacts were not reported because they were run only within the subset of Workers and Schoolers, respectively.

#### Interplay between setting and TU profiles

The association between TU profiles and contact numbers significantly changed across settings. In the rural setting, Schoolers reported the largest mean number of contacts, around 16 per day, which was 25% higher than what reported by Homestayers (Table S2).

Instead, in both urban settings, those spending time at work or farther from home in the general community were characterized by higher numbers of social contacts. In the slum, Workers reported 35% more contacts than Homestayers, while Walkers reported 37% more contacts (Table S3). Finally, in the mixed/non-slum setting, Workers and Commuters reported 70% and 68% more contacts than those Homestayers, corresponding to a mean of 10.3 and 11.4 daily contacts, respectively (Table S4).

**Table S2** Survey's participants and numbers of contacts per day (over the two survey days) in the rural setting (N=512): reported average numbers and relative numbers of contacts (incidence rate ratio, IRR) per day (from GEE model with negative binomial distribution and overdispersion parameter equal to 0.087, 95% CI [0.072, 0.11]).

| Variable | Category | Participants | | Number of contacts | | Relative number of contacts | |
|---|---|---|---|---|---|---|---|
| | | N | % | Mean | SD | IRR | 95% CI |
| Sex | Female | 276 | 53.9 | 11.50 | 0.28 | 1.00 | [1.00,1.00] |
| | Male | 236 | 46.1 | 11.63 | 0.37 | 0.99 | [0.92,1.06] |
| Age group | 00-01y | 36 | 7.0 | 7.46 | 0.46 | 0.74 | [0.62,0.88] |
| | 01-05y | 92 | 18.0 | 10.49 | 0.43 | 1.01 | [0.88,1.17] |
| | 06-14y | 136 | 26.6 | 13.38 | 0.38 | 1.13 | [1.01,1.28] |
| | 15-18y | 48 | 9.4 | 13.76 | 0.93 | 1.13 | [0.97,1.32] |
| | 19-34y | 91 | 17.8 | 11.21 | 0.54 | 1.00 | [1.00,1.00] |
| | 35-59y | 70 | 13.7 | 10.95 | 0.58 | 1.01 | [0.89,1.14] |
| | 60y+ | 39 | 7.6 | 8.38 | 0.52 | 0.86 | [0.74,1.00] |
| HH size | 1-2p | 31 | 6.1 | 7.43 | 0.69 | 1.00 | [1.00,1.00] |
| | 3p | 42 | 8.2 | 11.13 | 0.65 | 1.46 | [1.18,1.82] |
| | 4p | 54 | 10.5 | 10.98 | 0.67 | 1.46 | [1.15,1.85] |
| | 5-6p | 152 | 29.7 | 10.74 | 0.39 | 1.42 | [1.13,1.80] |
| | 7p+ | 233 | 45.5 | 12.93 | 0.33 | 1.70 | [1.35,2.15] |
| Living arrangements | One generation | 73 | 14.3 | 9.87 | 0.58 | 1.00 | [1.00,1.00] |
| | Two generations | 358 | 69.9 | 11.95 | 0.27 | 0.96 | [0.83,1.11] |
| | Three generations | 78 | 15.2 | 11.53 | 0.56 | 0.89 | [0.75,1.05] |
| | Missing | 3 | 0.6 | 10.19 | 1.03 | 0.76 | [0.62,0.94] |
| SES | Low | 370 | 72.3 | 11.30 | 0.27 | 1.00 | [1.00,1.00] |
| | Medium | 131 | 25.6 | 12.24 | 0.46 | 1.03 | [0.96,1.11] |
| | High | 11 | 2.1 | 12.01 | 1.03 | 1.16 | [0.93,1.44] |
| Education level | Primary | 252 | 49.2 | 12.56 | 0.36 | 1.00 | [1.00,1.00] |
| | Secondary | 31 | 6.1 | 11.40 | 0.61 | 0.95 | [0.84,1.09] |
| | College/university | 7 | 1.4 | 13.41 | 0.81 | 1.14 | [0.96,1.36] |
| | None | 222 | 43.4 | 10.07 | 0.28 | 0.88 | [0.80,0.96] |
| TU profile | Homestayers | 359 | 70.1 | 11.16 | 0.25 | 1.00 | [1.00,1.00] |
| | Workers | 13 | 2.5 | 10.94 | 1.15 | 1.03 | [0.85,1.25] |
| | Schoolers | 39 | 7.6 | 15.95 | 0.82 | 1.25 | [1.13,1.37] |
| | Walkers | 65 | 12.7 | 11.70 | 0.69 | 1.05 | [0.95,1.16] |
| | Commuters | 34 | 6.6 | 10.60 | 0.73 | 0.98 | [0.87,1.11] |
| | Travelers | 2 | 0.4 | 9.50 | 0.35 | 1.17 | [0.99,1.40] |
| Day type | Weekday | 327 | 63.9 | 11.40 | 0.25 | 1.00 | [1.00,1.00] |
| | Weekend | 185 | 36.1 | 11.95 | 0.40 | 1.11 | [1.04,1.19] |

**Table S3** Survey's participants and numbers of contacts per day (over the two survey days) in the urban slum setting (N=345): reported average numbers and relative numbers of contacts (incidence rate ratio, IRR) per day (from GEE model with negative binomial distribution and overdispersion parameter equal to 0.055, 95% CI [0.036, 0.083]).

| Variable | Category | Participants | | Number of contacts | | Relative number of contacts | |
|---|---|---|---|---|---|---|---|
| | | N | % | Mean | SD | IRR | 95% CI |
| Sex | Female | 217 | 62.9 | 6.95 | 0.25 | 1.00 | [1.00,1.00] |
| | Male | 128 | 37.1 | 6.37 | 0.28 | 0.93 | [0.84,1.03] |
| Age group | 00-01y | 39 | 11.3 | 4.75 | 0.37 | 0.68 | [0.55,0.83] |
| | 01-05y | 52 | 15.1 | 6.15 | 0.41 | 0.89 | [0.72,1.09] |
| | 06-14y | 45 | 13.0 | 8.35 | 0.52 | 1.09 | [0.92,1.29] |
| | 15-18y | 19 | 5.5 | 6.72 | 0.63 | 1.07 | [0.87,1.32] |
| | 19-34y | 128 | 37.1 | 6.55 | 0.27 | 1.00 | [1.00,1.00] |
| | 35-59y | 55 | 15.9 | 6.40 | 0.51 | 1.02 | [0.87,1.19] |
| | 60y+ | 7 | 2.0 | 6.35 | 0.73 | 1.10 | [0.84,1.44] |
| HH size | 1-2p | 80 | 23.2 | 5.57 | 0.34 | 1.00 | [1.00,1.00] |
| | 3p | 93 | 27.0 | 6.31 | 0.31 | 1.20 | [1.02,1.40] |
| | 4p | 73 | 21.2 | 6.95 | 0.43 | 1.31 | [1.07,1.62] |
| | 5-6p | 60 | 17.4 | 8.44 | 0.49 | 1.53 | [1.24,1.89] |
| | 7p+ | 18 | 5.2 | 7.85 | 0.64 | 1.39 | [1.07,1.81] |
| | Missing | 21 | 6.1 | 5.82 | 0.71 | 1.05 | [0.80,1.36] |
| Living arrangements | One generation | 149 | 43.2 | 5.91 | 0.26 | 1.00 | [1.00,1.00] |
| | Two generations | 173 | 50.1 | 7.43 | 0.27 | 1.01 | [0.87,1.18] |
| | Three generations | 2 | 0.6 | 9.21 | 2.26 | 1.80 | [1.29,2.50] |
| | Missing* | 21 | 6.1 | 5.82 | 0.71 | | |
| SES | Low | 67 | 19.4 | 6.81 | 0.43 | 1.00 | [1.00,1.00] |
| | Medium | 233 | 67.5 | 6.87 | 0.24 | 1.01 | [0.89,1.15] |
| | High | 45 | 13.0 | 5.91 | 0.42 | 0.82 | [0.69,0.96] |
| Education level | Primary | 134 | 38.8 | 7.44 | 0.32 | 1.00 | [1.00,1.00] |
| | Secondary | 57 | 16.5 | 6.00 | 0.37 | 0.91 | [0.79,1.05] |
| | College/university | 13 | 3.8 | 6.76 | 0.96 | 1.12 | [0.84,1.49] |
| | None | 123 | 35.7 | 6.28 | 0.29 | 1.01 | [0.87,1.18] |
| | Missing | 18 | 5.2 | 5.36 | 0.53 | 0.71 | [0.57,0.87] |
| TU profile | Homestayers | 233 | 67.5 | 6.38 | 0.22 | 1.00 | [1.00,1.00] |
| | Workers | 61 | 17.7 | 7.37 | 0.45 | 1.35 | [1.20,1.53] |
| | Schoolers | 23 | 6.7 | 7.70 | 0.49 | 1.13 | [0.96,1.31] |
| | Walkers | 14 | 4.1 | 8.88 | 0.87 | 1.37 | [1.18,1.58] |
| | Commuters | 11 | 3.2 | 5.66 | 0.75 | 0.97 | [0.79,1.20] |
| | Travelers | 2 | 0.6 | 10.69 | 1.87 | 1.79 | [1.26,2.53] |
| | Missing | 1 | 0.3 | 1.00 | 0.00 | 0.18 | [0.14,0.24] |
| Day type | Weekday | 208 | 60.3 | 6.95 | 0.23 | 1.00 | [1.00,1.00] |
| | Weekend | 137 | 39.7 | 6.36 | 0.26 | 0.90 | [0.84,0.96] |

* The category "Missing" for the variable "Living arrangements" is omitted from the GEE model for collinearity with the same category in the variable "HH size".

**Table S4** Survey's participants and numbers of contacts per day (over the two survey days) in the urban mixed/non-slum setting (N=550): reported average numbers and relative numbers of contacts (incidence rate ratio, IRR) per day (from GEE model with negative binomial distribution and overdispersion parameter equal to 0.21, 95% CI [0.18, 0.25]).

| Variable | Category | Participants | | Number of contacts | | Relative number of contacts | |
|---|---|---|---|---|---|---|---|
| | | N | % | Mean | SD | IRR | 95% CI |
| Sex | Female | 365 | 66.4 | 7.75 | 0.25 | 1.00 | [1.00,1.00] |
| | Male | 185 | 33.6 | 7.67 | 0.43 | 0.97 | [0.87,1.09] |
| Age group | 00-01y | 56 | 10.2 | 4.49 | 0.31 | 0.53 | [0.41,0.70] |
| | 01-05y | 48 | 8.7 | 7.86 | 0.63 | 0.89 | [0.67,1.16] |
| | 06-14y | 51 | 9.3 | 10.04 | 0.68 | 1.45 | [1.21,1.74] |
| | 15-18y | 72 | 13.1 | 8.60 | 0.52 | 1.19 | [1.00,1.41] |
| | 19-34y | 174 | 31.6 | 6.70 | 0.34 | 1.00 | [1.00,1.00] |
| | 35-59y | 123 | 22.4 | 8.03 | 0.51 | 1.11 | [0.96,1.28] |
| | 60y+ | 26 | 4.7 | 8.27 | 0.76 | 1.07 | [0.83,1.38] |
| HH size | 1-2p | 92 | 16.7 | 7.67 | 0.70 | 1.00 | [1.00,1.00] |
| | 3p | 115 | 20.9 | 6.59 | 0.44 | 0.99 | [0.80,1.21] |
| | 4p | 113 | 20.5 | 7.93 | 0.49 | 1.04 | [0.84,1.29] |
| | 5-6p | 152 | 27.6 | 7.96 | 0.39 | 1.08 | [0.87,1.33] |
| | 7p+ | 65 | 11.8 | 9.06 | 0.42 | 1.21 | [0.97,1.51] |
| | Missing | 13 | 2.4 | 6.12 | 0.90 | 0.77 | [0.36,1.65] |
| Living arrangements | One generation | 233 | 42.4 | 7.39 | 0.38 | 1.00 | [1.00,1.00] |
| | Two generations | 279 | 50.7 | 7.91 | 0.28 | 1.00 | [0.87,1.15] |
| | Three generations | 21 | 3.8 | 9.09 | 0.81 | 1.01 | [0.81,1.26] |
| | Missing | 17 | 3.1 | 7.17 | 1.50 | 1.33 | [0.65,2.71] |
| SES | Low | 9 | 1.6 | 7.43 | 1.50 | 1.00 | [1.00,1.00] |
| | Medium | 107 | 19.5 | 7.17 | 0.53 | 1.05 | [0.74,1.48] |
| | High | 434 | 78.9 | 7.85 | 0.25 | 1.09 | [0.79,1.52] |
| Education level | Primary | 112 | 20.4 | 8.27 | 0.45 | 1.00 | [1.00,1.00] |
| | Secondary | 186 | 33.8 | 8.30 | 0.39 | 1.15 | [0.99,1.33] |
| | College/university | 107 | 19.5 | 6.34 | 0.44 | 0.90 | [0.76,1.07] |
| | None | 138 | 25.1 | 7.82 | 0.48 | 1.44 | [1.14,1.81] |
| | Missing | 7 | 1.3 | 4.89 | 0.79 | 0.69 | [0.53,0.90] |
| TU profile | Homestayers | 403 | 73.3 | 7.02 | 0.22 | 1.00 | [1.00,1.00] |
| | Workers | 53 | 9.6 | 10.29 | 0.73 | 1.70 | [1.47,1.96] |
| | Schoolers | 42 | 7.6 | 9.23 | 0.81 | 1.30 | [1.10,1.53] |
| | Walkers | 29 | 5.3 | 8.12 | 0.91 | 1.07 | [0.89,1.28] |
| | Commuters | 18 | 3.3 | 11.40 | 1.60 | 1.68 | [1.37,2.07] |
| | Travelers | 4 | 0.7 | 4.04 | 1.17 | 0.86 | [0.61,1.23] |
| | Missing | 1 | 0.2 | 6.00 | 0.00 | 0.80 | [0.66,0.98] |
| Day type | Weekday | 373 | 67.8 | 7.87 | 0.28 | 1.00 | [1.00,1.00] |
| | Weekend | 177 | 32.2 | 7.44 | 0.27 | 0.99 | [0.93,1.06] |
